# A NOVEL AIR-DRIED MULTIPLEX HIGH RESOLUTION MELT ASSAY FOR THE DETECTION OF EXTENDED SPECTRUM BETA-LACTAMASE AND CARBAPENEMASE GENES

**DOI:** 10.1101/2021.02.18.21251950

**Authors:** Ana I. Cubas-Atienzar, Christopher T. Williams, Abhilasha Karkey, Sabina Dongol, Manandhar Sulochana, Shrestha Rajendra, Glyn Hobbs, Katie Evans, Patrick Musicha, Nicholas Feasey, Luis E. Cuevas, Emily R. Adams, Thomas Edwards

## Abstract

Here we describe the development and evaluation of a novel an air-dried high-resolution melt (HRM) assay to detect eight major extended spectrum beta-Lactamase (ESBL) (SHV and CTXM groups 1 and 9) and Carbapenemase (NDM, IMP, KPC, VIM and OXA-48) genes that cause antimicrobial resistance. The assay was evaluated using 440 DNA samples extracted from bacterial isolates from Nepal, Malawi and UK and 390 clinical Enterobacteriaceae isolates with known resistance phenotypes from Nepal. The sensitivity and specificity for detecting the ESBL and Carbapenemase genes in comparison to the reference gel-base PCR and sequencing was 94.7% (95%CI: 92.5%-96.5%) and 99.2% (95%CI: 98.8%-99.5%) and 98.5% (95%CI: 97.0%-99.4%) and 98.5% (95%CI: 98.0%-98.9%) when compared to the original wet format. The overall phenotypic agreement was 91.1% (95%CI: 90.0%-92.9%) on predicting resistance to cefotaxime and carbapenems. We observed good inter-machine reproducibility of the air-dried HRM assay using the Rotor-Gene Q, QuantStudio™ 5, CFX96, LightCycler^®^ 480 and MIC. Assay stability upon storage in the fridge (6.2°C ± 0.9), room temperature (20.35°C ± 0.7) and oven (29.7°C ± 1.4) were assessed at six time points for eight months and no loss of sensitivity occurred under all conditions. We present here a ready-to-use air-dried HRM-PCR assay that offers an easy, thermostable, fast and accurate tool for the detection of ESBL and Carbapenamase genes to improve AMR diagnosis and treatment.

## INTRODUCTION

Antimicrobial resistance (AMR) is a major global cause of death and the development of new antibiotics is considered a public health priority.^1^ AMR causes an estimated 700,000 deaths globally each year, and this number is predicted to rise to 10 million by 2050. ^2^ Identification of AMR is typically by culture-based phenotypic antimicrobial susceptibility testing (AST) which require incubation, from primary sample, for 48 to 96 hours. As clinical management decisions are often taken rapidly, the lack of timeliness of AST leads to an inaccurate diagnosis and inappropriate treatment. ^3,4^ First line or broad-spectrum antibiotics are often used in large doses to ensure their efficacy on the suspected but unknown aetiological pathogens.^3,5^ Empirical treatment facilitates the emergence of AMR, increases the duration of hospitalisation, damages the patient microbiota and increases the cost of therapy.^6–8^ Rapid diagnosis of AMR can enable targeted usage of antibiotics, improved patient outcomes and antimicrobial stewardship.^3,5^ Improved use of antibiotics through the development of rapid diagnostics is an important approach to tackle AMR.^9^

The most common mechanism of drug resistance in Gram-negative bacteria is mediated by the production of β-lactamases, including the extended-spectrum β-lactamases (ESBLs) and Carbapenemases,^10^ which provide resistance to the β-lactam antibiotics. Polymerase Chain Reaction (PCR) based detection of ESBLs and Carbapenemase genes may provide a faster diagnosis of AMR than phenotypic methods, which might in turn generate more timely information for treatment decisions.^11,12^ Whist molecular methods for the detection and characterisation of microbial resistance genes are becoming increasingly established, with good agreement with phenotypic methods, producing faster results, ^13–15^ their use in clinical settings is, however, hampered by the high degree of multiplexing needed due to the many genes involved in resistance. In addition, PCR requires a cold chain to maintain the integrity of reagents, equipment, and trained staff, which are often unavailable in low-resource settings, especially in low- and middle-income countries (LMICs). One approach that could facilitate the implementation of PCR assays in LMICs would be to provide the PCR primers, Taq enzyme and buffer components dry in the PCR plastics. This process would eliminate the need for the cold chain, ensure their biological integrity and simplify preparation, as only nuclease-free water and the DNA template would need to be added to resuspend the PCR reagents.^16–18^ Typically, this process would be done by lyophilisation of the reagents. Lyophilisation, also called freeze-drying, is the process of the removal of water from a product by volatilization and desorption to increase the lifespan of a product. However, lyophilisation process is costly and requires the addition of excipients, such as cryoprotectants and bulking agents.^17,19^

We report here the development and validation of an air dried HRM-PCR mix to detect the most frequent ESBLs and Carbapenemase enzymes based on a previously validated in-house AMR HRM-PCR assay.^20^

## METHODS

### Air-dried HRM assay optimisation

We adapted an in-house 9-plex HRM PCR that detects nine major ESBL genes (TEM, SHV and CTXM groups 1 and 9) and Carbapenemase genes (NDM, IMP, KPC, VIM and OXA-48) developed in our laboratory ^20^ to a dry format. For the dry-out process, AmpDRY™ (Biofortuna, UK) was used, which is a PCR reaction mix that allows direct air drying of the whole reaction including primers and reporter molecules and removes the need for lyophilisation systems and reagents. The composition of each HRM reaction included a mixture of 1x EvaGreen® dye (Biotium, Canada), primers for detecting ESBL groups and Carbapenemase genes, ^20^ the proprietary air-drying PCR buffer AmpDRY™ (Biofortuna, UK) and PCR grade water to a final volume of 6.25μl. The reaction mixture was added into each of the wells of a 96-well PCR plate (Starlab, Germany) and was dried in an oven-drier (ElextriQ, UK) at 35°C for 17 hours. PCR was performed by adding 2.5μl of bacterial DNA and 500 mM Betaine (Sigma Aldrich, UK) in PCR grade water to each PCR well containing the dried reagents for a final reaction volume of 12.5μl. PCR plates were briefly centrifugated before PCR amplification and, when plates were not compatible with the thermocycler used, the mixture was transferred to the appropriate reaction vessels. The optimised PCR amplification protocol consisted of an initial incubation step at 80°C for 15 minutes, followed by 30 cycles of denaturation for 10 seconds at 95°C, annealing for 60 seconds at 66°C and elongation for 10 seconds at 72°C monitoring the fluorescence in the FAM/SYBR channel. HRM analysis was carried out over a temperature range of 75°C to 95°C taking a reading in the HRM/SYBR channel every 0.1°C, with a 2 second stabilisation between each step. Positivity was indicated by a peak at the predictive melting temperature (Tm) of the target visualised as the negative first derivative of the melting curve. The Rotor-Gene Q (Qiagen, UK) was used for all the experiments except where stated otherwise. Optimal conditions of the assay were achieved by titration of individual reaction components, optimisation of amplification conditions and drying time. The original primer mix and their concentrations were as previously described, however the TEM was removed. ^20^

### Stored bacterial DNA and reference molecular tests

A panel of 440 DNA samples from well documented multidrug resistant (MDR) bacterial isolates from Nepal (n=294), the UK (n=103) and Malawi (n=43) was used to optimise and evaluate the air-dried HRM assay.

#### DNA from Nepal

this comprises isolates collected from 2012 to 2016 at Patan Hospital and includes strains of *Escherichia coli* (n=112), *Acinetobacter* spp. (n=72), *Klebsiella pneumoniae* (n=59), *Enterobacter* spp. (n=34), *Pseudomonas aeruginosa* (n=10), *Klebsiella oxytoca* (n=4), *Proteus* spp. (n=1), *Providencia retgerii* (n=1), and *Serratia rubidaea* (n=1).

#### DNA from Malawi

isolates were collected between 1998 and 2016 at Queen Elizabeth Central Hospital and comprised *E. coli* (n=25) and *K. pneumoniae* (n=18). The collection of isolates was approved by the University of Malawi College of Medicine Research and Ethics Committee (COMREC), Blantyre, under study number P.08/14/1614.

#### DNA from the UK

isolates were collected between 2012 and 2017 from the UK National Health Service hospitals and included *E. coli* (n=40), *K. pneumoniae* (n=27), *Enterobacter aerogenes* (n=12), *Enterobacter cloacae* (n=13), *Citrobacter freundii* (n=4), *P. aeruginosa* (n=4), *Morganella morganii* (n=2), and *K. oxytoca* (n=1).

DNA from the Nepal and Malawi isolates was extracted using the boilate ^21^ method and isolates from the UK were extracted using the DNeasy Blood and Tissue kit (Qiagen). The isolates sourced in the UK and Nepal were screened for ESBL and Carbapenemase markers using reference PCR published protocols^11,12^ with some modifications and the air-dried HRM assay. The reference PCR reaction mix was performed using DreamTaq PCR reaction mix (Thermo Fisher, UK), 2.5μl of DNA and nuclease free water to a final volume of 12.5μl. PCR amplification was visualised with PicoGreen™ (Life Technologies, USA) staining on a 1% TBE (Tris-borate-EDTA) gel with 1% to 2% of agarose depending on the fragment size to resolve. This reference gel-based PCR was not performed with the Malawian isolates as Next Generation sequencing data was available from previous studies. ^7,20^ In addition, the 440 isolates were screened using the in-house 9-plex HRM PCR assay originally developed in our laboratory ^20^ using the commercially available Type-it® HRM kit (Qiagen).

### Bacterial strains for phenotype prediction evaluation from Nepal

A set of 390 Gram negative bacteria with known phenotypes were chosen based on their resistance to imipenem (34%), meropenem (37%) and cefotaxime (85%) from a collection of characterised clinical isolates banked at Patan Hospital in Nepal. Isolates included strains of *E. coli* (n=72), *K. pneumoniae* (n=107), *Acinetobacter* spp. (n=73), *Enterobacter* (n=63), *K. oxytoca* (n=16), *P. aeruginosa* (n=13), *M. morganii* (n=3), *P. rettgeri* (n=1), *Proteus* spp. (n=2), *Serratia* spp. (n=3), *Salmonella* Typhi (n=25) *and Salmonella* Paratyphi (n=7). Isolates were resuscitated on MacConkey or nutrient agar and DNA extracted by a boiling lysis method as described elsewhere. ^21^

### Limit of detection

Limit of detection (LOD) of the air-dried assay was evaluated for the ESBL genes CTXM-1 and SHV, one positive for CTXM-1 (isolate 1), one positive for SHV (isolate 2), and a positive isolate for both genes (isolate 3) to estimate the LOD in isolates coproducing multiple genes. Two aliquots of 200μl of each of the suspensions were taken and processed following two extraction methodologies: DNeasy Blood and Tissue kit (Qiagen) and the boilate technique. DNA samples for each dilution series were tested in triplicate using the HRM assay. The LOD was defined as the lowest concentration at which the AMR genes were detected in all three replicates.

### Cross-platform validation

To evaluate the compatibility of the air-dried HRM assay in a wide range of platforms, a set of 94 samples comprising all the resistance genes were tested using different qPCR systems including the Rotor-Gene Q, QuantStudio™ 5 (Thermofisher, USA), CFX96 (BioRad, USA), LightCycler^®^ 480 (Roche Life Sciences, Germany) and MIC (Bio Molecular Systems, Australia). Amplification of the markers was assessed together with changes in Tms between platforms.

### Evaluation of the stability upon storage at different temperatures

Stability of the dried-HRM assay was evaluated over time under different storge temperatures. A set of 89 samples comprising all the markers and isolates 1-3 at the dilution of the LOD and previous dilution were tested with plates stored at different conditions. One PCR plate with the dried reaction mix was stored for each of the following periods of time; one week (T1), two weeks (T2), one (T3), three (T4) and eight months (T5) and at fridge (5°C), room (20°C) and oven temperature (30°C). PCR plates were sealed with foil adhesive film and individually wrapped in heat sealed aluminium foil laminated pouches containing one desiccant sachet (Merck, USA). Temperature and humidity were recorded weekly.

### Data analysis

Statistical evaluations were performed with SPSS v.19 (2010, US). The outcome of all tests was labelled as 0 when negative or 1 when positive. The level of agreement between tests was determined using Cohen’s Kappa. Kappa coefficients (κ) with values between 0 and 0.20, 0.21 and 0.39, 0.40 and 0.59, 0.60 and 0.79, 0.80 and 0.90 and 91 to 1 were interpreted as no agreement, minimal, weak, moderate, strong, and almost perfect agreement, respectively. ^22^ Statistical significance of differences in Tms between platforms was measured using One-Way-ANOVA and differences of peak height between different storage conditions using One-Way-ANOVA with Tukey’s test for Post-Hoc analysis. Statistical significance was set at a p-value < 0.05.

## Results

### Air-dried HRM assay evaluation using banked DNA

The air-dried HRM assay was capable of identifying the eight markers, each of which was characterised by the presence of a single peak at the expected Tm (Fig. 1a). The assay was also able to identify isolates co-producers of four AMR markers (Fig. 1b). There was no overlap between adjacent peaks with a minimum separation of peak Tm of 0.8 °C allowing easy identification of multiple genes within the same sample.

**Figure 1.**
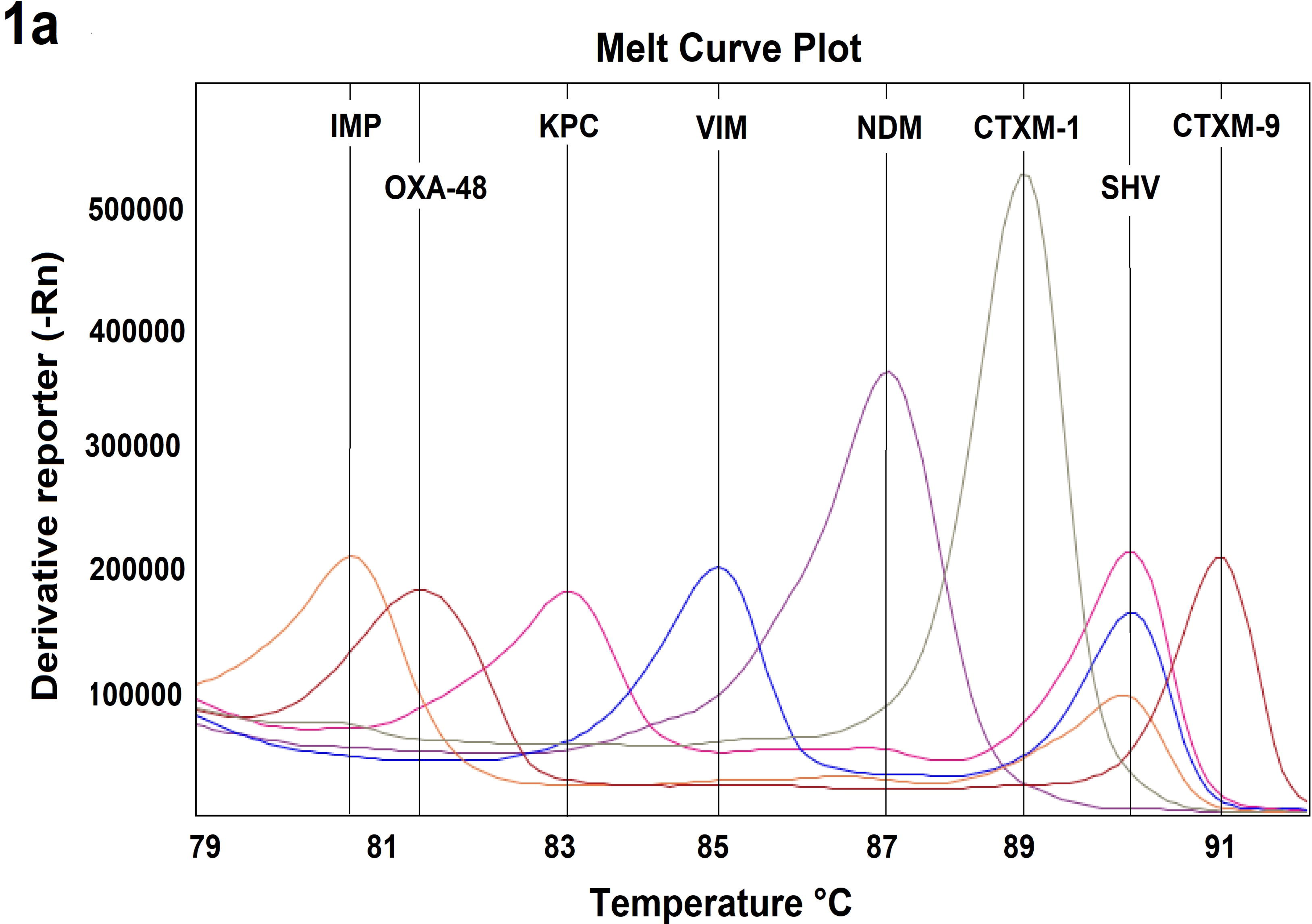
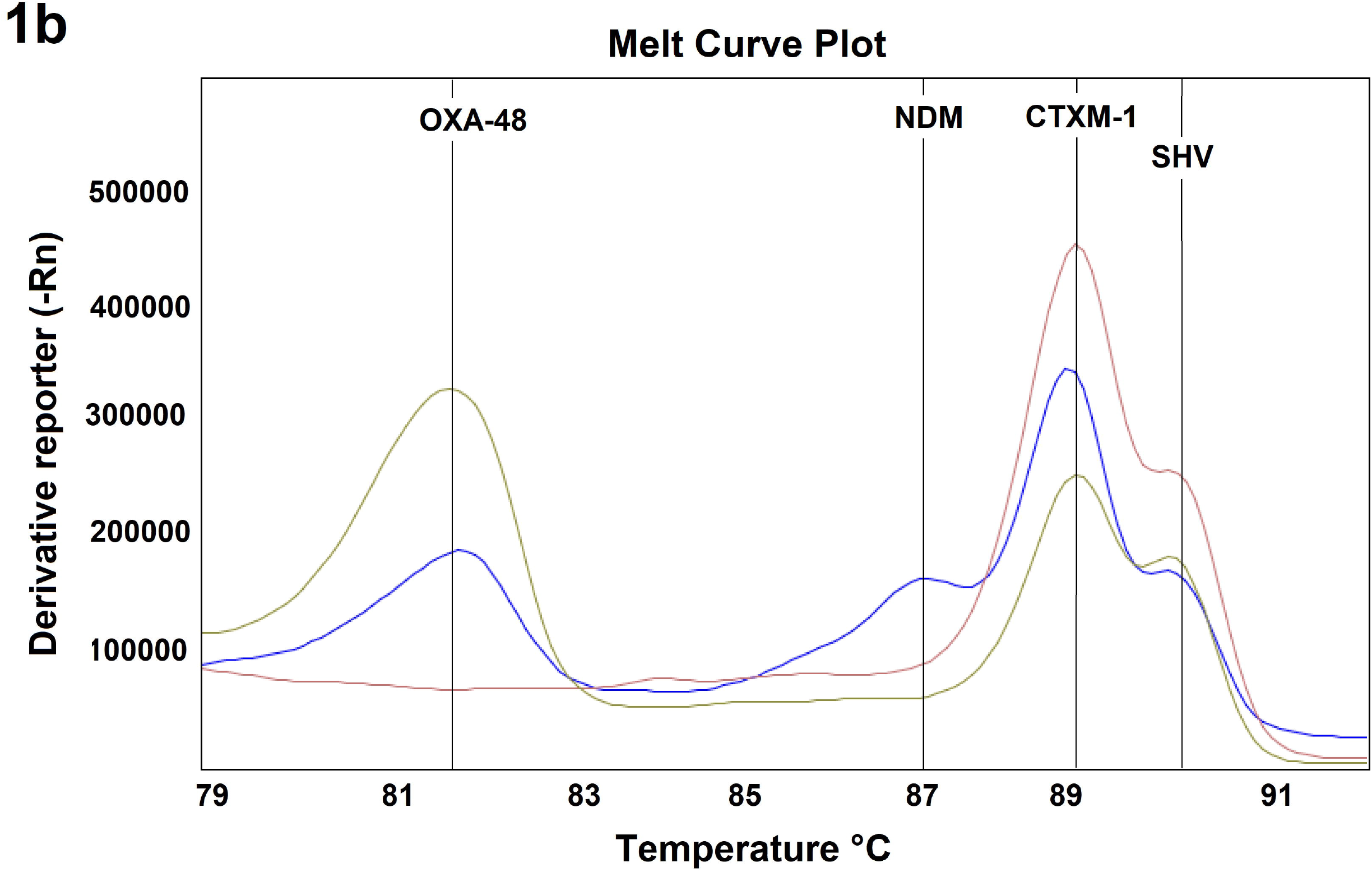
Melt curve profile of the air-dried HRM assay showing, 1a) the panel comprising the eight markers and, 1b) simultaneous detection of two (pink), three (yellow) and four genes (blue) in isolates coproducers of ESBL and carbapenemases genes.

Measures of diagnostic accuracy and agreement of the air-dried HRM assay for detecting individual genes compared to the reference tests are detailed in Table 1 (PCR and WGS) and Table 2 (original 9-Plex HRM assay). The overall sensitivity and specificity of the air-dried HRM assay for all genetic markers in comparison with the reference gel-based PCR and sequencing were 94.7% (95%CI: 92.5%-96.5%) and 99.2% (95%CI: 98.8%-99.5%) and, in comparison with the original 9-plex HRM PCR assay ^20^ were 98.5% (95%CI: 97.0%-99.4%) and 98.5% (95%CI: 98.0%-98.9%). When compared with the reference gel-based PCR, the air-dried HRM assay had almost perfect agreement (κ = 0.94-1) for the ESBL CTXM group and Carbapenemase markers and moderate agreement (κ = 0.79) for SHV. SHV was often found in coproducers of multiple genes and the sensitivity of SHV was lower in isolates carrying two (76.7%) and three genes (59.3%) than single producers of SHV (92.6%).

**Table 1.** Sensitivity (S), specificity (SP), accuracy (ACC) and agreement (k) of the air-dried HRM assay for detecting individual genes compared to the reference PCR and WGS.

| | Reference PCR/WGS | | S (95% CI) | SP (95% CI) | ACC (95% CI) | $\kappa$ |
| --- | --- | --- | --- | --- | --- | --- |
|  | Positives | Negatives |  |  |  |  |
| CTXM-1 |  |  |  |  |  |  |
| Positives | 242 | 10 | 99.2% | 94.9% | 97.3% | 0.94 |
| Negatives | 2 | 185 | 97.7%-100% | 91.7%-97.5% | 95.3%-98.6% |  |
| CTXM-9 |  |  |  |  |  |  |
| Positives | 14 | 1 | 100% | 99.8% | 99.8% | 0.96 |
| Negatives | 0 | 422 | 76.8%-100% | 98.3%-99.6% | 98.7%- 99.9% |  |
| SHV |  |  |  |  |  |  |
| Positives | 94 | 8 | 79.7% | 97.5% | 92.7% | 0.79 |
| Negatives | 24 | 314 | 71.3%-86.5% | 95.2%-98.9% | 89.9% --95% |  |
| NDM |  |  |  |  |  |  |
| Positives | 112 | 3 | 99.1% | 99.7% | 99.1% | 0.98 |
| Negatives | 1 | 321 | 90.2%-98.6% | 98.3%-100% | 97%-99.5% |  |
| IMP |  |  |  |  |  |  |
| Positives | 2 | 0 | 100% | 100% | 100% | 1.00 |
| Negatives | 0 | 438 | 15.8% -100% | 99.2%-100% | 99.2%-100% |  |
| KPC |  |  |  |  |  |  |
| Positives | 8 | 0 | 100% | 100% | 100% | 1.00 |
| Negatives | 0 | 432 | 63.1%-100% | 99%-100% | 99.2% -100% |  |
| OXA-48 |  |  |  |  |  |  |
| Positives | 13 | 0 | 92.9% | 100% | 99.8% | 0.96 |
| Negatives | 1 | 426 | 66.1%-99.8% | 99.2%-100% | 98.8% - 100% |  |
| VIM |  |  |  |  |  |  |
| Positives | 17 | 2 | 100% | 99.5% | 99.6% | 0.94 |
| Negatives | 0 | 421 | 80.5%-100% | 98.3%-99.9% | 98.4%-99.9% |  |

**Table 2.**
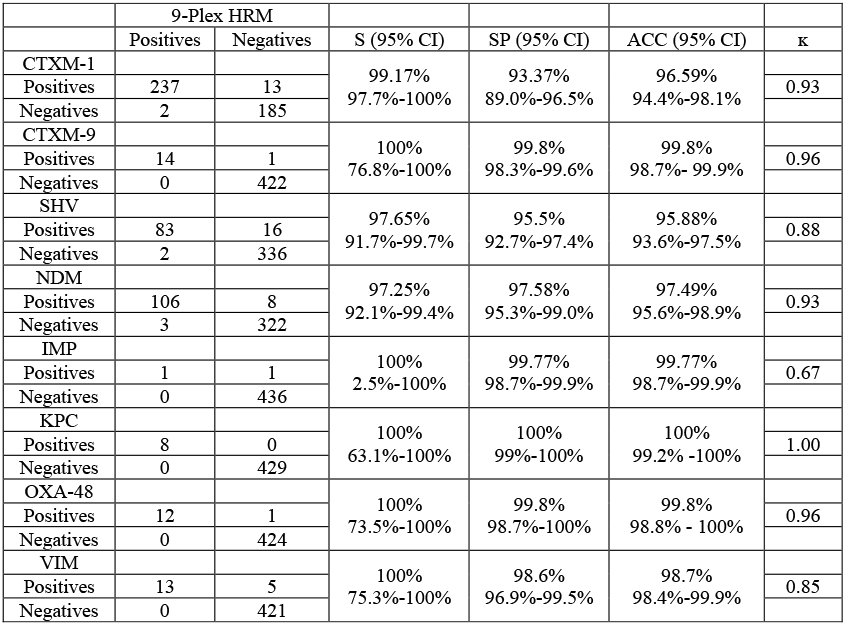
Sensitivity (S), specificity (SP), accuracy (ACC) and agreement (k) of the air-dried HRM for detecting individual genes compared to the original 9-Plex HRM assay^20^ using Type-it^®^ HRM buffer (Qiagen).

### Bacterial strains for phenotype prediction evaluation from Nepal

The overall agreement of the air-dried HRM result and phenotype was 91.1% (95%CI: 89.0%-92.9%) for Enterobacteriaceae isolates and 59.0% (95%CI: 52.8%-64.1%) for non-Enterobacteriaceae isolates (*Acinetobacter* spp., *P. aeruginosa* and *H. influenzae*). The air-dried HRM assay had strong agreement with the phenotype (κ = 0.820) among Enterobacteriaceae isolates with a sensitivity on predicting resistance to cefotaxime of 92.7% (95%CI: 88.9%-95.4%) and on predicting resistance to carbapenems 83.9% (95%CI: 76.2%-87.9%). However, the phenotype was poorly predicted among non-Enterobacteriaceae isolates using the air-dried HRM assay (κ = 0.251).

### Cross-platform validation

A good reproducibility was obtained on all instruments. Peak calling was performed by visual observation by the presence of a peak at the expected Tm and cut-off was established for each instrument by evaluating five threshold values set as 20%, 10%, 7.5%, 5% and 3% of the fluorescence of the highest peak. The optimal cut-off for the Rotor-Gene Q, QuantStudio and MIC was 5% of the fluoresce of the highest peak and for CFX96 and LightCycler® 480 it was 10%. These cut-offs produced almost perfect agreement with the reference tests (κ =0.935).

The amplicon Tm (°C) shifted across platforms (Fig. 2) and ranged from ± 0.013°C to ± 0.99°C for CTXM-1, ± 0.07-1.09°C for CTXM-9, ± 0.08-1.15°C for IMP, ±0.02-1.26°C for KPC, ±0.01-1.38°C for NDM, ±0.19-1.5°C for OXA-48, ±0.08-0.94°C for SHV and ±0.12-1.27°C depending on the platform used. The Tm differences within the same peak and neighbouring peaks is shown in Tables 3a and 3b for each of the platforms. The Tm difference was not statistically significant for any of the platforms for either the type of peak, peaks within the same cluster (p=0.318) and neighbouring clusters (p=1.00).

**Table 3.**
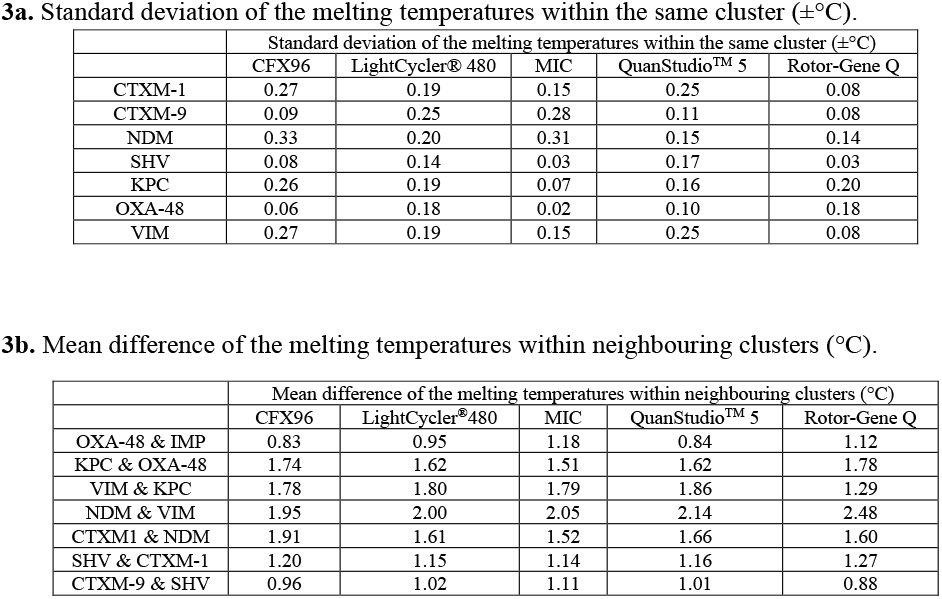
Variability of melting temperature within the same and between neighbouring cluster obtained in the validated platforms.

**3a.** Standard deviation of the melting temperatures within the same cluster ( $\pm^{\circ}\text{C}$ ).
| | Standard deviation of the melting temperatures within the same cluster ($\pm^{\circ}\text{C}$) | | | | |
| --- | --- | --- | --- | --- | --- |
|  | CFX96 | LightCycler® 480 | MIC | QuanStudio™ 5 | Rotor-Gene Q |
| CTXM-1 | 0.27 | 0.19 | 0.15 | 0.25 | 0.08 |
| CTXM-9 | 0.09 | 0.25 | 0.28 | 0.11 | 0.08 |
| NDM | 0.33 | 0.20 | 0.31 | 0.15 | 0.14 |
| SHV | 0.08 | 0.14 | 0.03 | 0.17 | 0.03 |
| KPC | 0.26 | 0.19 | 0.07 | 0.16 | 0.20 |
| OXA-48 | 0.06 | 0.18 | 0.02 | 0.10 | 0.18 |
| VIM | 0.27 | 0.19 | 0.15 | 0.25 | 0.08 |

**3b.** Mean difference of the melting temperatures within neighbouring clusters ( $^{\circ}\text{C}$ ).
| | Mean difference of the melting temperatures within neighbouring clusters ($^{\circ}\text{C}$) | | | | |
| --- | --- | --- | --- | --- | --- |
|  | CFX96 | LightCycler®480 | MIC | QuanStudio™ 5 | Rotor-Gene Q |
| OXA-48 & IMP | 0.83 | 0.95 | 1.18 | 0.84 | 1.12 |
| KPC & OXA-48 | 1.74 | 1.62 | 1.51 | 1.62 | 1.78 |
| VIM & KPC | 1.78 | 1.80 | 1.79 | 1.86 | 1.29 |
| NDM & VIM | 1.95 | 2.00 | 2.05 | 2.14 | 2.48 |
| CTXM1 & NDM | 1.91 | 1.61 | 1.52 | 1.66 | 1.60 |
| SHV & CTXM-1 | 1.20 | 1.15 | 1.14 | 1.16 | 1.27 |
| CTXM-9 & SHV | 0.96 | 1.02 | 1.11 | 1.01 | 0.88 |

**Figure 2.**
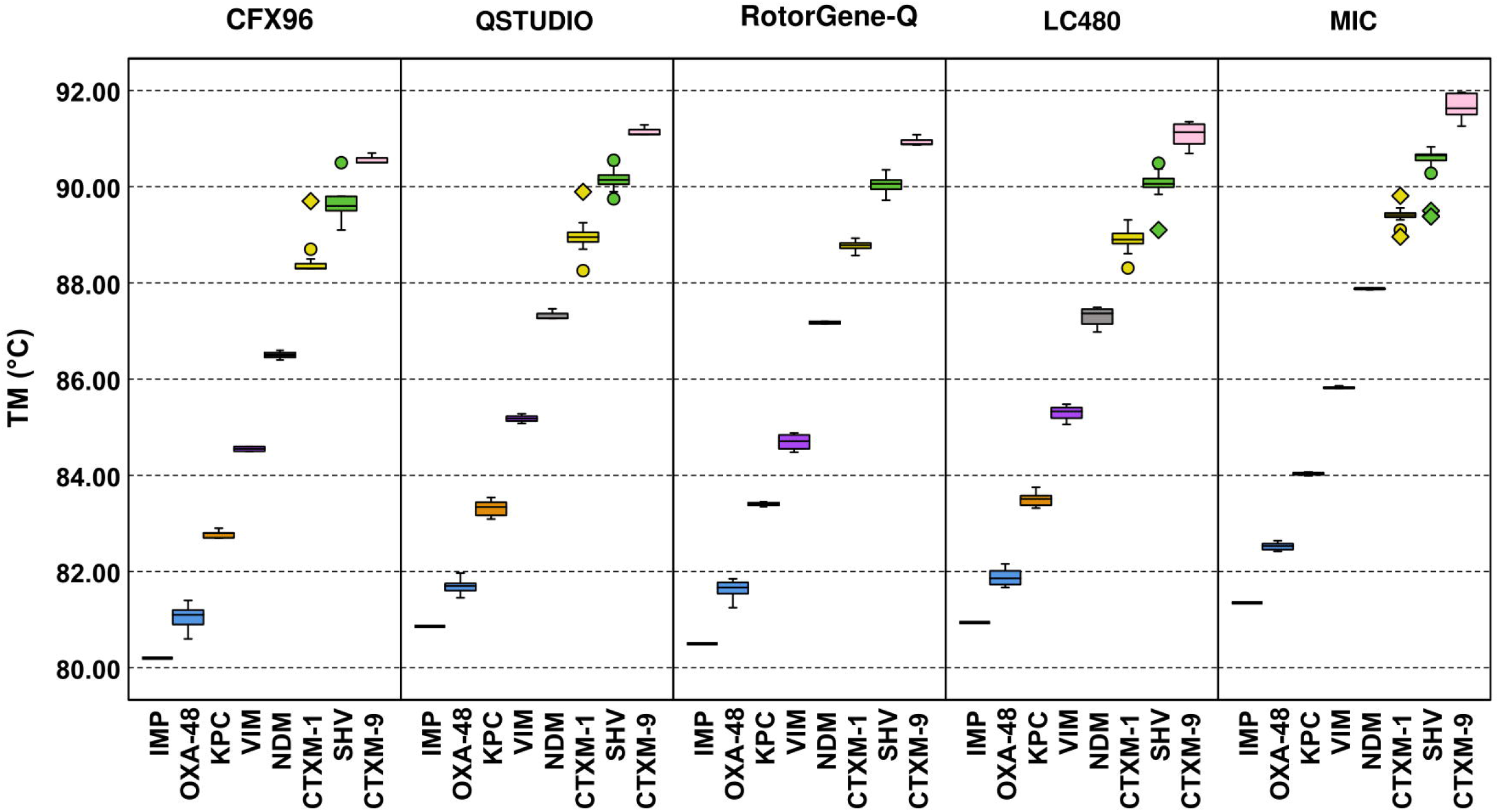
Melting temperatures of the eight amplicons of the air-dried HRM assay ran in the CFX96, QuanStudioTM 5 (QStudio), Rotor-Gene-Q (RotorGene-Q), LightCycler® 480 (LC48) and MIC.The whiskers show the maximum and minimum values, with the exceptions of outliers (circles) and extremes (rhombus).

### Limit of detection

The limit of detection was 11.5, 102 and 960 cfu/reaction using DNeasy kit and 2.3, 20.4 and 192 cfu/reaction by the boilate method for isolates carrying the CTXM-1, SHV and both CTXM-1 and SHV genes, respectively.

### Stability upon different storage conditions

The effect of storage time and temperature was assessed on the AmpDry mix by analysing the plate mean fluorescence peak height and amplification of isolates, including isolates at LOD dilutions. The average temperature for room storage, fridge and oven was 20.35°C ± 0.7, 6.2°C ± 0.9 and 29.7°C ± 1.4 respectively, the humidity of the room was at 36.5% ± 9.34. Overall, room temperature was the best storage condition compared to fridge and oven. The difference of mean fluoresce peak hight was not statistically significant within the same time point but was statistically significant between different time points (Fig. 3). The peak height started decreasing after storage time T3 for room and oven storage, and at T2 for fridge storage (Fig. 3). Nonetheless, the difference of mean peak height produced with the AmpDry mix stored at time T3 (one month) was not statistically significant to the produced at T0, T1 and T2 at all storage conditions. The AmpDry mix recovered at T4 and T5 (fridge only) produced significantly lower peak heights when compared to T1, T2 and T3 (room temperature only). The mean peak height produced with the AmpDry mix stored at time T5 at room temperature, was comparable to all time points at all storage conditions and timepoints except at T1 for fridge storage (Fig. 3).

**Figure 3.**
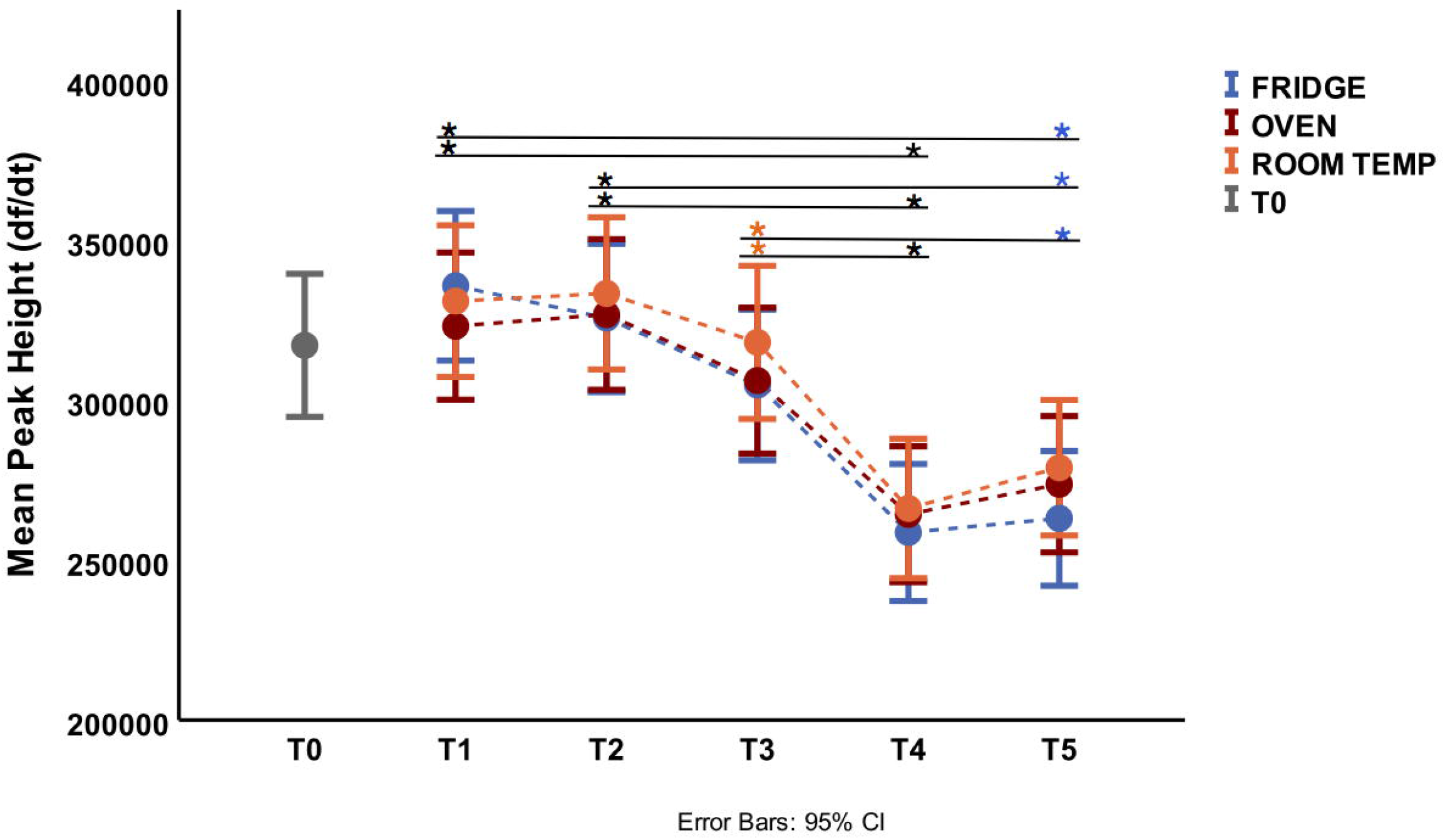
Plate mean fluoresce peak height at the beginning of study (T0), one week (T1), two weeks (T2), one month (T3), three months (T4) and eight months (T5) under fridge storage (6.2°C ± 0.9), room temperature (20.35°C ± 0.7) and oven (29.7°C ± 1.4). Colour of asterisks indicates which storage conditions were statistically different between time points: blue (fridge), orange (room temperature), red (oven), black (all temperature conditions).

Isolate 1 was negative at the LOD dilution at T3 under oven storage; isolate 2 was negative at the LOD dilution at T3 under room temperature and oven storage, and isolate 3 was positive in all runs tested (Fig 4). Of the 89 isolates tested, 100% were positive for all markers at all storage times and conditions, except for one sample that had one of three marker peaks below the cut-off (NDM) at T4 fridge storage (data not shown).

**Figure 4.**
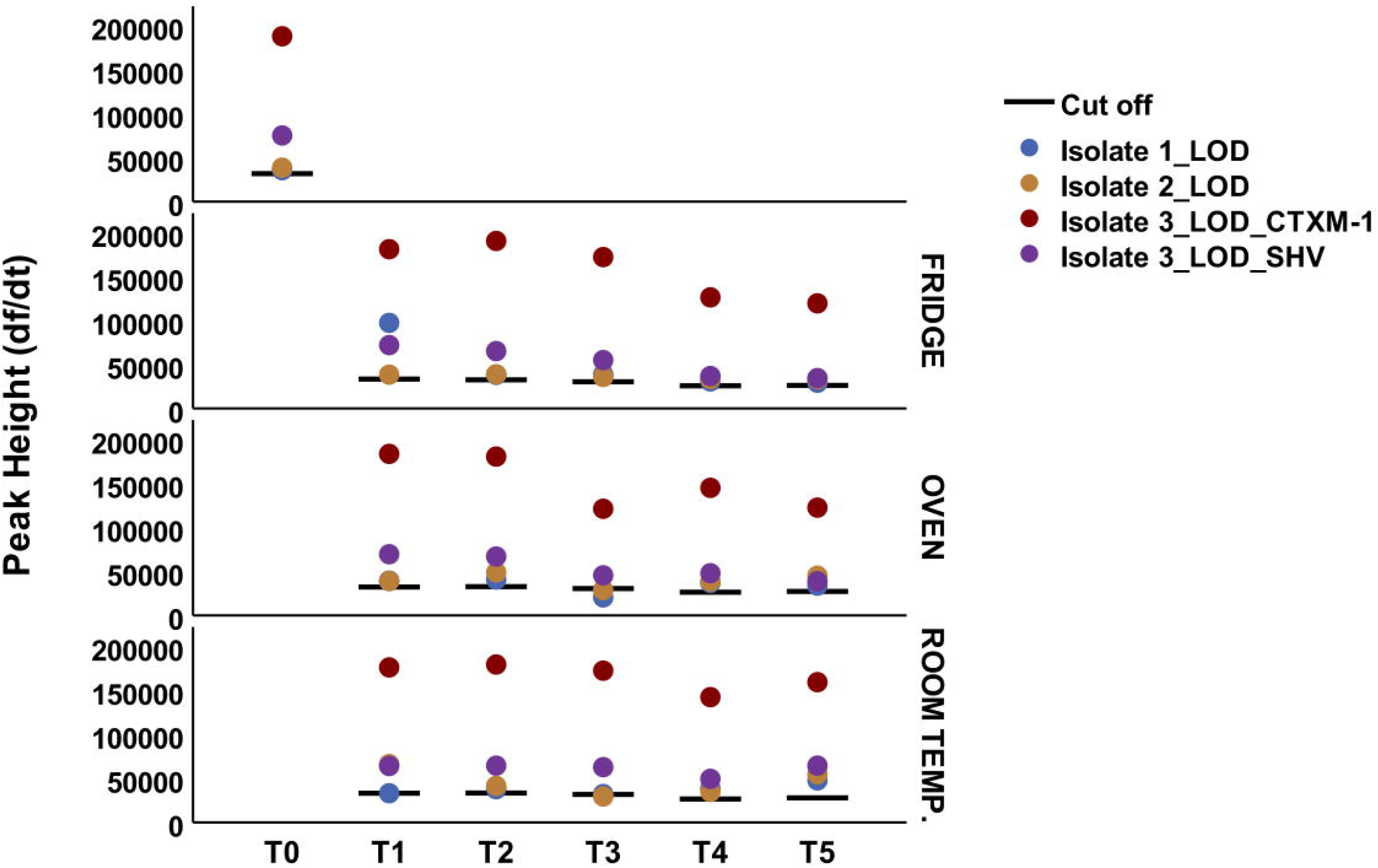
Peak height of the isolates 1 (CTX-M-1 positive), 2 (SHV positive) and 3 (CTX-M-1 and SHV positive) at LOD dilution at different timepoints and storage conditions.

## Discussion

In this study, we evaluated the performance of an 8-plex HRM PCR assay in dry format to detect ESBL and Carbapenemase genes. The assay showed high sensitivity, specificity and measures of agreement for all markers when compared to the reference tests. In addition, the drying process did not result in loss of performance, with all the resistance genes of the 89 clinical isolates correctly classified after 6 months of storage.

The dry format of the assay overcomes key real-world challenges relating to transport, storage, and freezing/thawing issues, which can substantially lower the sensitivity of PCR.^23,24^ This HRM assay presents several major advantages over fresh qPCR mixes as its resistant to long periods of storage at relatively warm temperatures (30 °C) and its stability during handling in warm conditions enables easy storage and transportation of the assay at ambient temperature for long periods of time. This would be of particular importance in LMICs where laboratories face insufficient and suboptimal cold chain capacity.^25^

As this assay is easy to set-up and interpretation of results with analysis of the melting data can be automated, it may be straight-forward to implement in laboratories with access to qPCR facilities, but otherwise moderate resources, as all that is required is to reconstitute the mix and add template DNA. The assay has good performance using the boilate extraction method, which is fast, simple and easy to implement with minimal resources.

This is timely as global capacity in molecular diagnostics has recently surged in response to the COVID-19 pandemic. The level of multiplexing enables detection of the 8 major Carbapenemase and ESBL gene families in a single tube with a sensitivity and specificity compared to reference molecular tests, which require a panel of PCR assays. Molecular detection of AMR genes can provide useful epidemiological data and enable the tracking of particular resistance genes at a hospital or national level.^26^

Cross-platform validation illustrates a remarkably good performance on all 5 q-PCR systems (Rotor-Gene Q, QuantStudio™ 5, CFX96, LightCycler^®^ 480 and MIC) evaluated, with minimal variation on the peak Tms, which was not statistically significant. The cut-offs however required slight adjustment (5% or 10% of the highest peak) to achieve the best performance, nevertheless this is straightforward correction that can be applied with simple instructions.

The protocol has some constraints as a 24h incubation from primary sample to grow the isolates is still required prior DNA extraction. The assay has not been evaluated using direct clinical samples but the LOD obtained here indicates sensitivity to be insufficient to detect the low CFU/ml (>1/ml) possible in bacterial bloodstream infections. ^27,28^

The overall agreement to predict bacterial phenotypes was strong (κ = 0.82) amongst Enterobacteriaceae isolates but weak in non-Enterobacterial isolates. Thus, we do not recommend the use of the assay in non-Enterobacterial isolates. The high discrepancy among non-Enterobacteriaceae isolates can be explained as *Acinetobacter* spp. and *Pseudomas* spp. have many other mechanisms of resistance such as efflux pumps, chromosomal mediated AmpC enzymes, permeability defects, and modifications of target sites that are less common in the family Enterobacteriaceae. ^29,30^ Possible causes of false negative results amongst Enterobacteriaceae isolates include the carriage of less common β-lactamase genes that are not covered by the HRM assay, such as plasmid mediated AmpC enzymes^31^, or GES-1^32^. Other reasons for phenotype-genotype mismatches include enzyme modifications that change the spectrum of activity and susceptibility profile ^33^, and also isolates with MICs close to the breakpoint being incorrectly classified during phenotypic susceptibility testing. ^34^

To summarise, the air-dried HRM assay detected ESBL and Carbapenemese genes fast, effectively and with high specificity and sensitivity and maintained performance after six months of storage at room temperatures. This 8-plex dry HRM assay was also successfully transferred to 5 different PCR platforms indicating that can be reliably implemented in many laboratories. The assay can become a useful tool for AMR diagnosis and surveillance.

## Data Availability

All data is available on request from the corresponding author

## Funding

The study was funded through the MRC Proximity to Discovery (P2D) award number MC_PC_17196. The funders had no role in the design of the study, data collection, analysis, or preparation of the manuscript.

## Acknowledgments

We would like to thank the staff at Patan Hospital in Kathmandu for their assistance with the isolates used in this study and Biofortuna for advice.

## Authors’ contributions

A.I.C.A., T.E., E.R.A. and L.E.C. contributed to the conception and design of the study; A.I.C.A., C.W., S.M. and R.S. carried out the experimental work. A.I.C.A., T.E. and C.W. analysed the data. A.I.C.A. wrote the first draft of the manuscript. All authors were involved in the manuscript preparation and revision, and approval of the final version of the manuscript.

## Notes

### Competing Interest Statement

The authors have declared no competing interest.

### Author Declarations

University of Malawi College of Medicine Research and Ethics Committee (COMREC), Blantyre, under study number P.08/14/1614

